# Background rates of all-cause mortality, hospitalizations, and emergency department visits among nursing home residents in Ontario, Canada to inform COVID-19 vaccine safety assessments

**DOI:** 10.1101/2021.03.17.21253290

**Authors:** Maria Sundaram, Sharifa Nasreen, Andrew Calzavara, Siyi He, Hannah Chung, Susan E. Bronskill, Sarah A. Buchan, Mina Tadrous, Peter Tanuseputro, Kumanan Wilson, Sarah Wilson, Jeffrey C. Kwong, on behalf of the Canadian Immunization Research Network (CIRN) investigators

## Abstract

**Background:** Nursing home (NH) residents are prioritized for COVID-19 vaccination. We report monthly mortality, hospitalizations, and emergency department (ED) visit incidence rates (IRs) during 2010-2020 to provide context for COVID-19 vaccine safety assessments.

**Methods:** We observed outcomes among NH residents using administrative databases. IRs were calculated by month, sex, and age group. Comparisons between months were assessed using one-sample t-tests; comparisons by age and sex were assessed using chi-squared tests.

**Results:** From 2010-2019, there were 83,453 (SD: 652.4) NH residents per month, with an average of 2.3 (SD: 0.28) deaths, 3.1 (SD: 0.16) hospitalizations, and 3.6 (SD: 0.17) ED visits per 100 residents per month. From March to December 2020, mortality IRs were increased, but hospitalization and ED visit IRs were reduced (p<0.05).

**Conclusion:** We identified consistent monthly mortality, hospitalization, and ED visit IRs during 2010-2019. Marked differences in these rates were observed during 2020, coinciding with the COVID-19 pandemic.

## Introduction^1^

Nursing home (NH) residents have been disproportionately affected by the coronavirus disease 2019 (COVID-19) pandemic, comprising a substantial proportion of total COVID-19 deaths in many countries, including over 80% of COVID-19 deaths in Canada during the first epidemic wave [1, 2]. The vaccine rollout has prioritized, among others, NH residents because of their high risk of poor outcomes from COVID-19 [3].

However, NH residents are generally frail and are at comparatively higher risk of poor outcomes, even outside the context of a global pandemic [4, 5]. As a result, deaths and healthcare use within the expected range of event numbers for NH residents could create an erroneous impression that vaccine rollout is associated with increased risk of such events [6]. There is a lack of data on poor outcome rates among NH residents in Canada to facilitate comparisons of observed versus expected outcome rates during vaccination rollout. Therefore, we sought to quantify background rates of all-cause mortality, hospitalizations, and emergency department (ED) visits among all NH residents in Ontario, the most populous province in Canada. Since COVID-19 and related restrictions may also impact these outcomes, we report these rates separately during 2020.

## Methods

We conducted an observational analysis using population-level health administrative databases encompassing all residents of Ontario, Canada (2020 population: approximately 14.7 million, of which 10% live in rural areas; median age: 40.4 years) [7, 8]. Ontario has a single-payer health system that provides universal access to physician and hospital services for all Ontario residents, including NH residents, who receive personal and nursing care and subsidized housing [9]. In Ontario, NH may be not-for-profit (managed either by public, municipal, or private companies), or for-profit institutions managed by private companies [9].

These datasets were linked using unique encoded identifiers and analyzed at ICES (formerly called the Institute for Clinical Evaluative Sciences). ICES is a not-for-profit research institute that is a prescribed entity under Ontario’s Personal Health Information Protection Act (PHIPA). Section 45 of PHIPA authorizes ICES to collect personal health information, without consent, for the purpose of analysis or compiling statistical information with respect to the management of, evaluation, or monitoring of, the allocation of resources to or planning for all or part of the health system.

### Cohort definitions

We identified all Ontarians aged 18 years or older living in NHs using the physician billing database Ontario Health Insurance Plan (encompassing most outpatient interactions with the healthcare system), Ontario Drug Benefit (encompassing prescriptions), and the Continuing Care Reporting System (encompassing information about individuals in NH) databases. Information on age, sex and death were collected from the Registered Persons Database. We identified individual cohorts beginning at the start of each month from January 1, 2010 to December 31, 2019 inclusive, and from January 1, 2020 to December 31, 2020. Individuals were included in these cohorts if they were alive at the beginning of a month and recorded as living in a NH at least one day in that month or in the previous 90 days. We defined NH residents in this way because administrative datasets in Ontario may have incomplete capture of individual entry in, and exit from, NH care; but interactions with the healthcare system that would identify the individual as a NH resident are expected to happen at least once every 90 days. As a result, ICES uses a multi-component definition of NH resident that incorporates information from healthcare system interactions in a 90-day period that can identify individuals as NH residents.

### Statistical analysis

We calculated monthly rates per 100 NH residents overall, by sex, and by age group (18-69 years, 70-79 years, 80-89 years, and ≥90 years). We also conducted analyses comparing adults aged ≥80 years with adults younger than 80 years. Estimates from 2010-2019 were combined over years by month, and by sex and age group separately. Direct comparisons were conducted either with one-sample t-tests, in the case of comparisons by month from 2010-2019 vs. 2020, or with chi-squared tests, in the case of comparisons by sex or age. We also calculated the relative change in mortality in 2020, compared with mortality in 2010-2019, using a simple ratio (mortality in 2020 divided by average mortality in 2010-2019). As the first presumed SARS-CoV-2 positive case was identified in Canada on January 25, 2020, and Health Canada did not report that the primary source of infections was “local” transmission until March 24, 2020 [10], we defined the pandemic period in 2020 as beginning approximately on March 1, 2020.

## Results

From January 1, 2010 through December 31, 2019, there were an average of 83,453 (SD: 652.4) NH residents in Ontario in any given month (**Figure 1A**). Of these, an average of 57,104 (SD: 291.6; ∼68% of the total in any given month) were women, and an average of 58,733 (SD: 550.6; ∼70% of the total in any given month) were aged ≥80 years.

**Figure 1.**
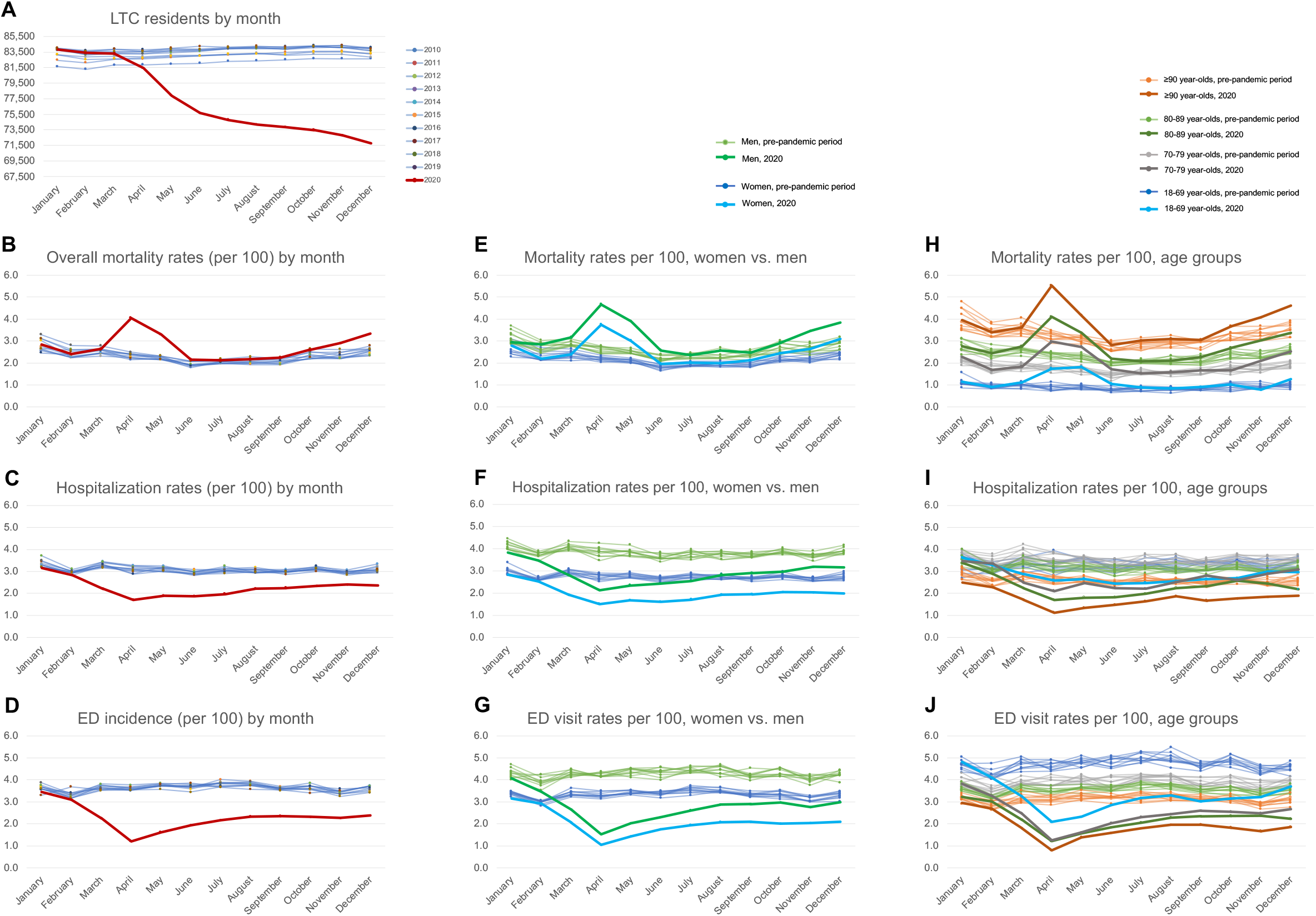
Nursing home residents (A) and monthly incidence rates of mortality, hospitalization, and emergency department visits per 100 nursing home residents (panels B-D), by sex (panels E-G) and by age (panels H-J).

During this period, approximately 2.3 (SD: 0.28) deaths per 100 NH residents occurred per month (**Figure 1B; Table 1**). These rates were higher for men (incidence rate [IR]: 2.6; SD: 0.31) than women (IR: 2.2; SD: 0.27) (p<0.001; **Figure 1E**), and for residents aged ≥80 years (IR: 5.6; SD: 0.68) than residents aged <80 years (IR: 2.6; SD: 0.33) (p<0.001; **Figure 1H**). An average of 0.4 (SD: 0.04) more monthly deaths per 100 residents occurred during winter months (December, January, February, and March) than during other months during 2010-2019 (p<0.001; **Table 1**).

**Table 1.**
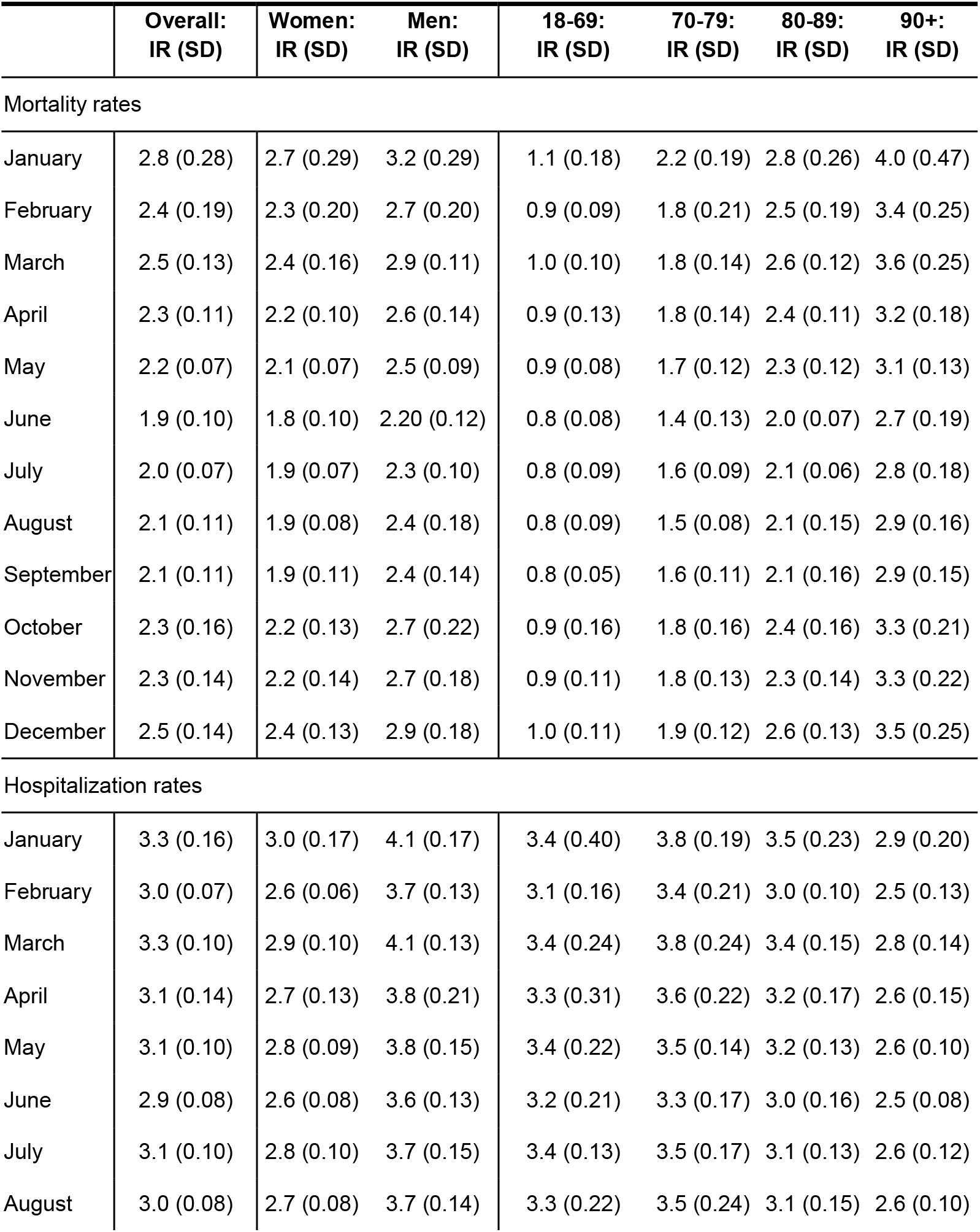
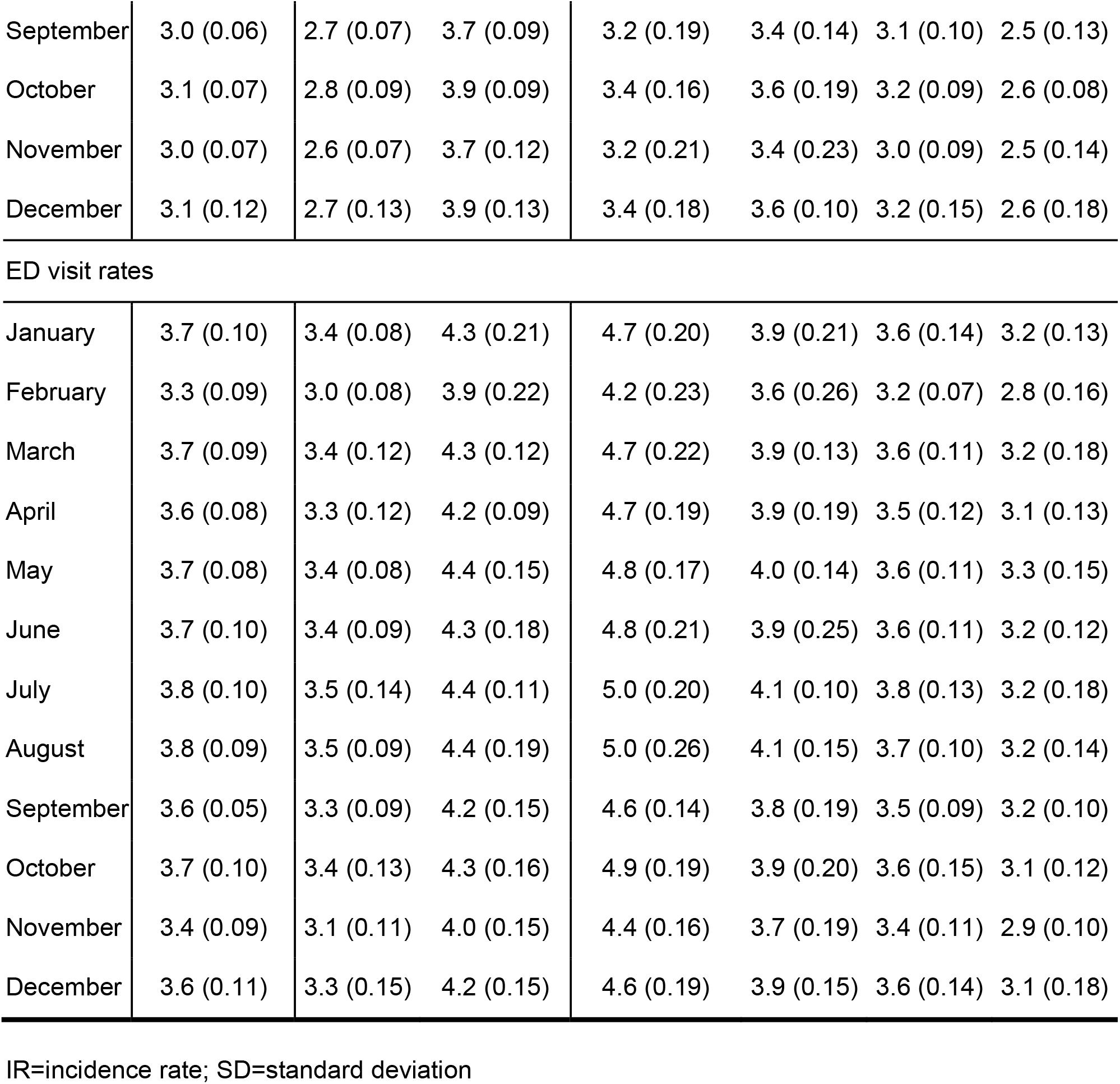
Mean incidence rates per 100 nursing home residents, monthly from 2010-2019.

|  | **Overall:**<br>**IR (SD)** | **Women:**<br>**IR (SD)** | **Men:**<br>**IR (SD)** | **18-69:**<br>**IR (SD)** | **70-79:**<br>**IR (SD)** | **80-89:**<br>**IR (SD)** | **90+:**<br>**IR (SD)** |
| --- | --- | --- | --- | --- | --- | --- | --- |
| Mortality rates |  |  |  |  |  |  |  |
| January | 2.8 (0.28) | 2.7 (0.29) | 3.2 (0.29) | 1.1 (0.18) | 2.2 (0.19) | 2.8 (0.26) | 4.0 (0.47) |
| February | 2.4 (0.19) | 2.3 (0.20) | 2.7 (0.20) | 0.9 (0.09) | 1.8 (0.21) | 2.5 (0.19) | 3.4 (0.25) |
| March | 2.5 (0.13) | 2.4 (0.16) | 2.9 (0.11) | 1.0 (0.10) | 1.8 (0.14) | 2.6 (0.12) | 3.6 (0.25) |
| April | 2.3 (0.11) | 2.2 (0.10) | 2.6 (0.14) | 0.9 (0.13) | 1.8 (0.14) | 2.4 (0.11) | 3.2 (0.18) |
| May | 2.2 (0.07) | 2.1 (0.07) | 2.5 (0.09) | 0.9 (0.08) | 1.7 (0.12) | 2.3 (0.12) | 3.1 (0.13) |
| June | 1.9 (0.10) | 1.8 (0.10) | 2.20 (0.12) | 0.8 (0.08) | 1.4 (0.13) | 2.0 (0.07) | 2.7 (0.19) |
| July | 2.0 (0.07) | 1.9 (0.07) | 2.3 (0.10) | 0.8 (0.09) | 1.6 (0.09) | 2.1 (0.06) | 2.8 (0.18) |
| August | 2.1 (0.11) | 1.9 (0.08) | 2.4 (0.18) | 0.8 (0.09) | 1.5 (0.08) | 2.1 (0.15) | 2.9 (0.16) |
| September | 2.1 (0.11) | 1.9 (0.11) | 2.4 (0.14) | 0.8 (0.05) | 1.6 (0.11) | 2.1 (0.16) | 2.9 (0.15) |
| October | 2.3 (0.16) | 2.2 (0.13) | 2.7 (0.22) | 0.9 (0.16) | 1.8 (0.16) | 2.4 (0.16) | 3.3 (0.21) |
| November | 2.3 (0.14) | 2.2 (0.14) | 2.7 (0.18) | 0.9 (0.11) | 1.8 (0.13) | 2.3 (0.14) | 3.3 (0.22) |
| December | 2.5 (0.14) | 2.4 (0.13) | 2.9 (0.18) | 1.0 (0.11) | 1.9 (0.12) | 2.6 (0.13) | 3.5 (0.25) |
| Hospitalization rates |  |  |  |  |  |  |  |
| January | 3.3 (0.16) | 3.0 (0.17) | 4.1 (0.17) | 3.4 (0.40) | 3.8 (0.19) | 3.5 (0.23) | 2.9 (0.20) |
| February | 3.0 (0.07) | 2.6 (0.06) | 3.7 (0.13) | 3.1 (0.16) | 3.4 (0.21) | 3.0 (0.10) | 2.5 (0.13) |
| March | 3.3 (0.10) | 2.9 (0.10) | 4.1 (0.13) | 3.4 (0.24) | 3.8 (0.24) | 3.4 (0.15) | 2.8 (0.14) |
| April | 3.1 (0.14) | 2.7 (0.13) | 3.8 (0.21) | 3.3 (0.31) | 3.6 (0.22) | 3.2 (0.17) | 2.6 (0.15) |
| May | 3.1 (0.10) | 2.8 (0.09) | 3.8 (0.15) | 3.4 (0.22) | 3.5 (0.14) | 3.2 (0.13) | 2.6 (0.10) |
| June | 2.9 (0.08) | 2.6 (0.08) | 3.6 (0.13) | 3.2 (0.21) | 3.3 (0.17) | 3.0 (0.16) | 2.5 (0.08) |
| July | 3.1 (0.10) | 2.8 (0.10) | 3.7 (0.15) | 3.4 (0.13) | 3.5 (0.17) | 3.1 (0.13) | 2.6 (0.12) |
| August | 3.0 (0.08) | 2.7 (0.08) | 3.7 (0.14) | 3.3 (0.22) | 3.5 (0.24) | 3.1 (0.15) | 2.6 (0.10) |
| September | 3.0 (0.06) | 2.7 (0.07) | 3.7 (0.09) | 3.2 (0.19) | 3.4 (0.14) | 3.1 (0.10) | 2.5 (0.13) |
| October | 3.1 (0.07) | 2.8 (0.09) | 3.9 (0.09) | 3.4 (0.16) | 3.6 (0.19) | 3.2 (0.09) | 2.6 (0.08) |
| November | 3.0 (0.07) | 2.6 (0.07) | 3.7 (0.12) | 3.2 (0.21) | 3.4 (0.23) | 3.0 (0.09) | 2.5 (0.14) |
| December | 3.1 (0.12) | 2.7 (0.13) | 3.9 (0.13) | 3.4 (0.18) | 3.6 (0.10) | 3.2 (0.15) | 2.6 (0.18) |
| ED visit rates |  |  |  |  |  |  |  |
| January | 3.7 (0.10) | 3.4 (0.08) | 4.3 (0.21) | 4.7 (0.20) | 3.9 (0.21) | 3.6 (0.14) | 3.2 (0.13) |
| February | 3.3 (0.09) | 3.0 (0.08) | 3.9 (0.22) | 4.2 (0.23) | 3.6 (0.26) | 3.2 (0.07) | 2.8 (0.16) |
| March | 3.7 (0.09) | 3.4 (0.12) | 4.3 (0.12) | 4.7 (0.22) | 3.9 (0.13) | 3.6 (0.11) | 3.2 (0.18) |
| April | 3.6 (0.08) | 3.3 (0.12) | 4.2 (0.09) | 4.7 (0.19) | 3.9 (0.19) | 3.5 (0.12) | 3.1 (0.13) |
| May | 3.7 (0.08) | 3.4 (0.08) | 4.4 (0.15) | 4.8 (0.17) | 4.0 (0.14) | 3.6 (0.11) | 3.3 (0.15) |
| June | 3.7 (0.10) | 3.4 (0.09) | 4.3 (0.18) | 4.8 (0.21) | 3.9 (0.25) | 3.6 (0.11) | 3.2 (0.12) |
| July | 3.8 (0.10) | 3.5 (0.14) | 4.4 (0.11) | 5.0 (0.20) | 4.1 (0.10) | 3.8 (0.13) | 3.2 (0.18) |
| August | 3.8 (0.09) | 3.5 (0.09) | 4.4 (0.19) | 5.0 (0.26) | 4.1 (0.15) | 3.7 (0.10) | 3.2 (0.14) |
| September | 3.6 (0.05) | 3.3 (0.09) | 4.2 (0.15) | 4.6 (0.14) | 3.8 (0.19) | 3.5 (0.09) | 3.2 (0.10) |
| October | 3.7 (0.10) | 3.4 (0.13) | 4.3 (0.16) | 4.9 (0.19) | 3.9 (0.20) | 3.6 (0.15) | 3.1 (0.12) |
| November | 3.4 (0.09) | 3.1 (0.11) | 4.0 (0.15) | 4.4 (0.16) | 3.7 (0.19) | 3.4 (0.11) | 2.9 (0.10) |
| December | 3.6 (0.11) | 3.3 (0.15) | 4.2 (0.15) | 4.6 (0.19) | 3.9 (0.15) | 3.6 (0.14) | 3.1 (0.18) |

IR=incidence rate; SD=standard deviation

We observed monthly averages of 3.1 (SD: 0.16) hospitalizations per 100 residents and 3.6 (SD: 0.17) ED visits per 100 residents during 2010-2019 (**Figures 1C and 1D; Table 1**). Hospitalization rates were higher for men (IR: 3.8; SD: 0.20) than women (IR: 2.7; SD: 0.15) (p<0.001) (**Figure 1F; Table 1**) and were lower for older residents (IR: 5.8; SD: 0.34) than younger residents (IR: 6.8; SD: 0.35) (p<0.001; **Figure 1I**). Similarly, ED visit rates were higher for men (IR: 4.3; SD: 0.21) than women (IR: 3.3; SD: 0.18) (p<0.001; **Figure 1G**) and lower for older residents (IR: 6.7; SD: 0.33) than younger residents (IR: 8.6; SD: 0.45) (p<0.001; **Figure 1J**).

We observed a steady decline in the total number of NH residents during 2020 (non-parametric test for trend p<0.01; **Figure 1A**). No statistically significant differences were observed in mortality rates in January or February of 2020 compared to those months in previous years. Mortality rates rose during March 2020, and were higher in all months of the pandemic period (March – December 2020) compared to corresponding months in 2010-2019 (p<0.05 for each month) (**Figure 1B, Table 2**); these differences were most pronounced in April and May 2020. Mortality was higher for men (**Figure 1E**) and older residents (**Figure 1H**). The ratio of excess deaths in 2020 to average mortality incidence in 2010-2019 was highest in April and May 2020, across sexes and age groups (**Table 3**).

**Table 2.**
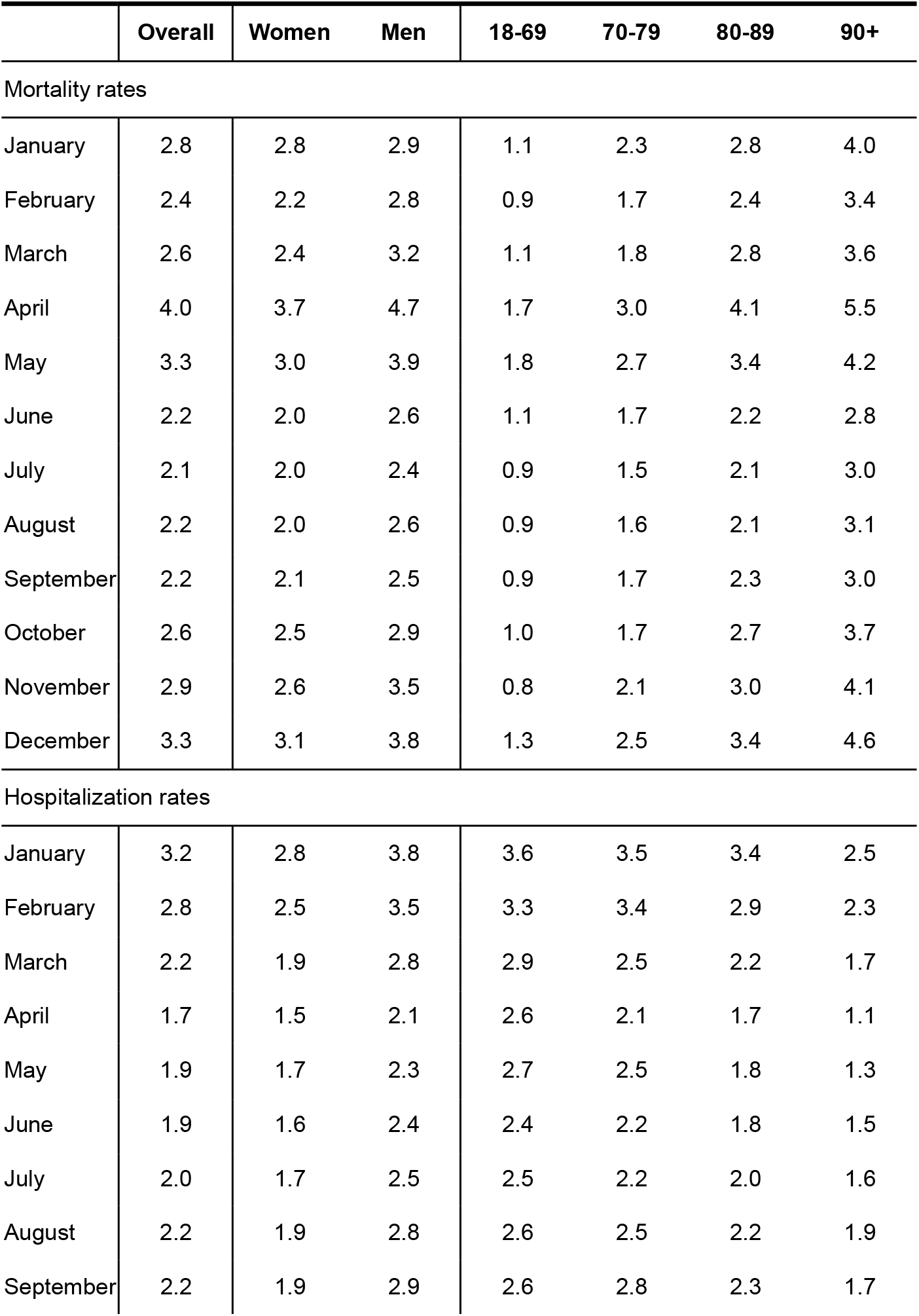
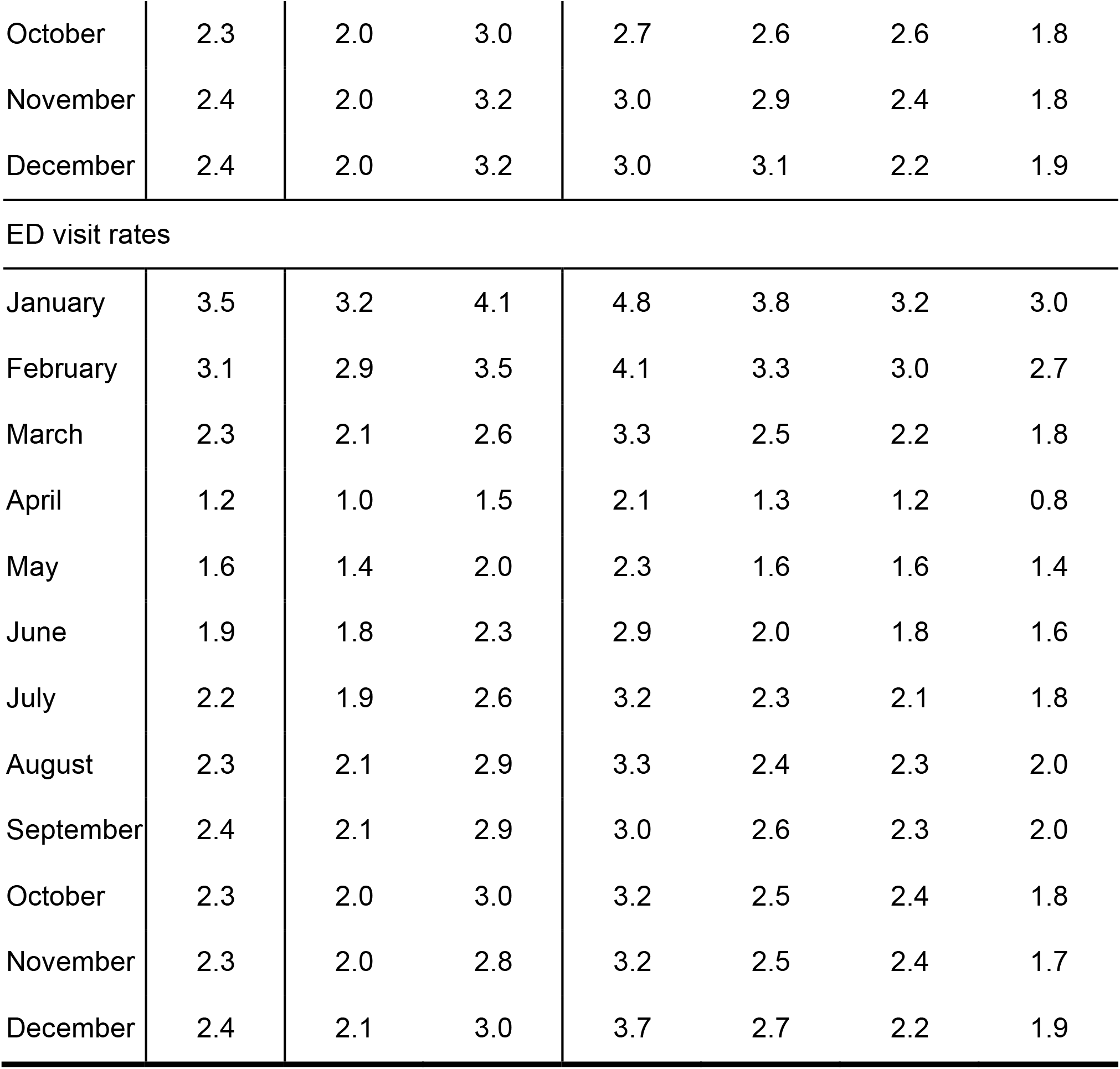
Incidence rates per 100 nursing home residents during 2020.

|  | Overall | Women | Men | 18-69 | 70-79 | 80-89 | 90+ |
| --- | --- | --- | --- | --- | --- | --- | --- |
| Mortality rates |  |  |  |  |  |  |  |
| January | 2.8 | 2.8 | 2.9 | 1.1 | 2.3 | 2.8 | 4.0 |
| February | 2.4 | 2.2 | 2.8 | 0.9 | 1.7 | 2.4 | 3.4 |
| March | 2.6 | 2.4 | 3.2 | 1.1 | 1.8 | 2.8 | 3.6 |
| April | 4.0 | 3.7 | 4.7 | 1.7 | 3.0 | 4.1 | 5.5 |
| May | 3.3 | 3.0 | 3.9 | 1.8 | 2.7 | 3.4 | 4.2 |
| June | 2.2 | 2.0 | 2.6 | 1.1 | 1.7 | 2.2 | 2.8 |
| July | 2.1 | 2.0 | 2.4 | 0.9 | 1.5 | 2.1 | 3.0 |
| August | 2.2 | 2.0 | 2.6 | 0.9 | 1.6 | 2.1 | 3.1 |
| September | 2.2 | 2.1 | 2.5 | 0.9 | 1.7 | 2.3 | 3.0 |
| October | 2.6 | 2.5 | 2.9 | 1.0 | 1.7 | 2.7 | 3.7 |
| November | 2.9 | 2.6 | 3.5 | 0.8 | 2.1 | 3.0 | 4.1 |
| December | 3.3 | 3.1 | 3.8 | 1.3 | 2.5 | 3.4 | 4.6 |
| Hospitalization rates |  |  |  |  |  |  |  |
| January | 3.2 | 2.8 | 3.8 | 3.6 | 3.5 | 3.4 | 2.5 |
| February | 2.8 | 2.5 | 3.5 | 3.3 | 3.4 | 2.9 | 2.3 |
| March | 2.2 | 1.9 | 2.8 | 2.9 | 2.5 | 2.2 | 1.7 |
| April | 1.7 | 1.5 | 2.1 | 2.6 | 2.1 | 1.7 | 1.1 |
| May | 1.9 | 1.7 | 2.3 | 2.7 | 2.5 | 1.8 | 1.3 |
| June | 1.9 | 1.6 | 2.4 | 2.4 | 2.2 | 1.8 | 1.5 |
| July | 2.0 | 1.7 | 2.5 | 2.5 | 2.2 | 2.0 | 1.6 |
| August | 2.2 | 1.9 | 2.8 | 2.6 | 2.5 | 2.2 | 1.9 |
| September | 2.2 | 1.9 | 2.9 | 2.6 | 2.8 | 2.3 | 1.7 |
| October | 2.3 | 2.0 | 3.0 | 2.7 | 2.6 | 2.6 | 1.8 |
| November | 2.4 | 2.0 | 3.2 | 3.0 | 2.9 | 2.4 | 1.8 |
| December | 2.4 | 2.0 | 3.2 | 3.0 | 3.1 | 2.2 | 1.9 |
| ED visit rates |  |  |  |  |  |  |  |
| January | 3.5 | 3.2 | 4.1 | 4.8 | 3.8 | 3.2 | 3.0 |
| February | 3.1 | 2.9 | 3.5 | 4.1 | 3.3 | 3.0 | 2.7 |
| March | 2.3 | 2.1 | 2.6 | 3.3 | 2.5 | 2.2 | 1.8 |
| April | 1.2 | 1.0 | 1.5 | 2.1 | 1.3 | 1.2 | 0.8 |
| May | 1.6 | 1.4 | 2.0 | 2.3 | 1.6 | 1.6 | 1.4 |
| June | 1.9 | 1.8 | 2.3 | 2.9 | 2.0 | 1.8 | 1.6 |
| July | 2.2 | 1.9 | 2.6 | 3.2 | 2.3 | 2.1 | 1.8 |
| August | 2.3 | 2.1 | 2.9 | 3.3 | 2.4 | 2.3 | 2.0 |
| September | 2.4 | 2.1 | 2.9 | 3.0 | 2.6 | 2.3 | 2.0 |
| October | 2.3 | 2.0 | 3.0 | 3.2 | 2.5 | 2.4 | 1.8 |
| November | 2.3 | 2.0 | 2.8 | 3.2 | 2.5 | 2.4 | 1.7 |
| December | 2.4 | 2.1 | 3.0 | 3.7 | 2.7 | 2.2 | 1.9 |

**Table 3.** Ratio of excess deaths in 2020 to average mortality incidence in previous years for COVID-19.

|  | Overall | Women | Men | 18-69 | 70-79 | 80-89 | 90 |
| --- | --- | --- | --- | --- | --- | --- | --- |
| Mortality rates |  |  |  |  |  |  |  |
| January | 1.01 | 1.05 | 0.93 | 1.04 | 1.05 | 0.99 | 0.99 |
| February | 0.99 | 0.96 | 1.05 | 0.98 | 0.95 | 0.99 | 1.00 |
| March | 1.05 | 1.01 | 1.10 | 1.14 | 0.99 | 1.07 | 1.02 |
| April | 1.74 | 1.71 | 1.78 | 1.99 | 1.67 | 1.73 | 1.71 |
| May | 1.48 | 1.44 | 1.55 | 2.02 | 1.61 | 1.49 | 1.35 |
| June | 1.11 | 1.08 | 1.16 | 1.36 | 1.20 | 1.11 | 1.05 |
| July | 1.04 | 1.05 | 1.01 | 1.09 | 0.98 | 1.01 | 1.06 |
| August | 1.05 | 1.03 | 1.08 | 1.05 | 1.03 | 1.01 | 1.06 |
| September | 1.07 | 1.09 | 1.04 | 1.11 | 1.05 | 1.09 | 1.05 |
| October | 1.11 | 1.12 | 1.09 | 1.09 | 0.95 | 1.13 | 1.13 |
| November | 1.24 | 1.20 | 1.30 | 0.85 | 1.20 | 1.30 | 1.23 |
| December | 1.31 | 1.30 | 1.34 | 1.29 | 1.31 | 1.31 | 1.31 |

Estimates are calculated with the following formula: (incidence rate in 2020)/(average incidence rate in 2010-2019).

There was a substantial reduction in hospitalizations during 2020 compared to previous years (p<0.001 for each month during the pandemic period; **Figure 1C** and **Table 2**). Hospitalization rates were higher for men than women (p<0.001; **Figure 1F**) and lower with increasing age throughout the pandemic period (**Figure 1I**). Similar to hospitalizations, there was a substantial reduction in ED visits during 2020 compared to previous years (p<0.001 for each month during 2020; **Figure 1D** and **Table 2**). Patterns by sex and age were similar as for hospitalizations (**Figures 1G** and **1J**).

## Discussion

This report presents rates of all-cause mortality, hospitalizations, and ED visits in NH residents in Ontario, Canada during a 10-year pre-pandemic period (2010-2019) and during 2020. Mortality rates in 2010-2019 are similar to rates reported by others in Canada [4], with rates being higher in men compared to women, and higher with increasing age. As expected, mortality rates spiked during the early phase of the COVID-19 pandemic, with high mortality rates among all age groups and sexes in April and May of 2020, similar to overall estimates for the province of Ontario [11]. Although mortality rates returned to values closer to pre-pandemic rates in August 2020, they rose again in November and December 2020, in keeping with the timing of Ontario’s second wave [12].

The decrease in mortality rates during summer 2020 is likely multifactorial. First, changes in provincial-level testing guidelines may have enabled earlier identification of COVID-19 cases. On April 8, 2020, Ontario officially expanded its testing guidelines beyond symptomatic NH residents to all residents, including all newly admitted individuals and asymptomatic residents who had contact with a confirmed-positive case [13]. Earlier identification of COVID-19 cases may have contributed to a subsequent reduction in mortality by preventing potential outbreaks and by providing needed care to residents earlier in the course of their illness. Second, enhanced infection prevention measures were implemented in NHs province-wide.[14] Third, reduced mortality rates in the summer months likely reflect a reduction in community-level transmission of SARS-CoV-2 experienced throughout Ontario [11].

Our analysis additionally identified a decline in hospitalizations and ED visits during 2020, compared to 2010-2019. This finding may initially seem counterintuitive but, taken in context with simultaneous increases in NH setting mortality and decreases in the total number of NH residents, these reductions likely indicate a reduction in hospitalized deaths, which has been reported previously [15]. There may have also been an increased unwillingness to seek hospital-based care for non-COVID-19-related issues during the beginning of the first pandemic wave; future analyses should also consider the reasons noted for ED visits and hospitalizations to identify COVID-19-related vs. non-COVID-19-related incidents. Our analysis identified that older adults (≥90 years) had the lowest hospitalization and ED visit rates both in pre-pandemic and pandemic periods; and that younger adults (18-64 years) had the highest hospitalization and ED visits in both periods. We believe these patterns reflect different goals of care for different age groups and populations.

This analysis uses large-scale health administrative datasets, which are not yet currently updated to the end of December 2020 with vital statistics. As a result, we are only able to report data up to the end of 2020, an important limitation of this work. If feasible, we will update this report as more data become available.

We intend for these findings to be used to contextualize potential increases in adverse events of special interest (AESI) [16] during the COVID-19 vaccine rollout in NH residents. Whereas mortality, hospitalization, and ED visit rates were highly stable over 10 pre-pandemic years, the marked changes in rates observed during 2020 indicate that caution is needed in determining the appropriate baseline to use when interpreting rates of AESIs following the COVID-19 vaccine rollout. Existing investigations on mortality in elderly and frail individuals who received COVID-19 vaccines has identified that mortality rates were consistent with high background rates of death in this population [17]; our data will help contextualize Canada- and Ontario-specific rates in a similar way.

## Conclusions

This report identified relatively consistent monthly mortality, hospitalization, and ED visit rates for Ontario NT residents over a ten-year pre-pandemic period. During 2020, we identified a substantial increase in mortality rates during April and May, followed by a regression during summer 2020 and subsequent increase in November and December 2020. In contrast, hospitalization and ED visit rates declined markedly in 2020 and (as of the latest data available) have not yet returned to baseline levels. These results provide important context for future analyses to identify potential AESI in NH residents.

## Data Availability

The dataset from this study is held securely in coded form at ICES. While legal data sharing agreements between ICES and data providers (e.g., healthcare organizations and government) prohibit ICES from making the dataset publicly available, access may be granted to those who meet pre-specified criteria for confidential access, available at www.ices.on.ca/DAS. The full dataset creation plan and underlying analytic code are available from the authors upon request, understanding that the computer programs may rely upon coding templates or macros that are unique to ICES and are therefore either inaccessible or may require modification.

## Authors’ contributions

JCK conceived of the study design and oversaw the study. AC obtained the data and calculated incidence rates. MS conducted additional statistical analysis and drafted the manuscript. PT, SEB, KW, and MT provided methodological input. All authors interpreted the results, critically revised the manuscript, and have approved the final version for publication.

## ^1^Abbreviations

NH: nursing home
COVID-19: coronavirus disease 2019
ED: emergency department

